# Assessing Red Blood Cell Distribution Width in Vietnamese Heart Failure Patients: A Cross-Sectional Study

**DOI:** 10.1101/2024.03.17.24304437

**Authors:** Hai Nguyen Ngoc Dang, Thang Viet Luong, Mai Thi Thu Cao, Vinh Trung Bui, Thanh Thien Tran, Hung Minh Nguyen

## Abstract

Heart failure (HF) is a growing public health concern, with red cell distribution width (RDW) gaining recognition as a cost-effective marker for predicting HF onset and progression. This study, conducted from February 2022 to February 2024 at Hue Central Hospital, aimed to assess RDW levels in hospitalized Vietnamese HF patients and its predictive value for HF severity, involving a cross-sectional analysis of 351 patients categorized into HF and non-HF groups. HF patients exhibited higher median RDW levels (14.90% [13.70% - 17.00%]) compared to non-HF individuals (13.10% [12.23% - 13.78%]). RDW was higher in HF patients with left ventricular ejection fraction (LVEF) < 50% (15.30%) than LVEF ≥ 50% (14.70%). ROC curve analysis for acute heart failure (AHF) prediction yielded an AUC of 0.651 (p < 0.001), with a cutoff point of 13.85%, sensitivity of 86.05%, and specificity of 36.94%. Elevated RDW levels were associated with HF risk (OR: 1.844, p < 0.001) and AHF (OR: 3.612, p < 0.001). Multivariate analysis identified associations between RDW and hemoglobin (Hb) (β = 2.431, p = 0.040) and hematocrit (HCT) (β = −3.355, p = 0.007) in HF. RBC and RDW > 13.85% were independent risk factors for AHF. This suggests that RDW levels can serve as biomarkers for diagnosing HF and predicting its severity. Their accessibility and cost-effectiveness indicate the potential utility of RDW in managing HF, particularly in settings with limited resources.

## Introduction

Heart failure (HF) represents a complex and life-threatening syndrome characterized by significant morbidity and mortality, limited functional capacity and quality of life, and substantial economic burden [1]. Additionally, HF has emerged as an increasingly critical public health concern. Given the aging population and enhanced survival rates following acute myocardial infarction, these trends are expected to persist [2]. HF has been recognized as a global pandemic, affecting an estimated 64.3 million individuals worldwide in 2017 [3]. Despite therapeutic advancements, HF remains a primary contributor to morbidity and mortality on a global scale [4].

The early detection and accurate diagnosis of HF are crucial for optimizing the treatment and prognoses of patients with this condition. Currently, HF diagnosis relies primarily on echocardiography and patients’ presenting symptoms, yet there lacks a definitive prognostic indicator for mortality among HF patients [5]. Biomarkers have shown promise in enhancing predictive capabilities alongside clinical evaluation in chronic heart failure (CHF) patients, although the incremental predictive value of individual biomarkers like B-type natriuretic peptide or troponin I in acute heart failure (AHF) is limited [6]. However, in Vietnam, the costs associated with these tests are prohibitive, and they are only available in large medical centers, presenting a significant challenge to their widespread application.

Red cell distribution width (RDW) is a parameter that reflects the variability in red blood cell size (anisocytosis), easily obtainable from a complete blood count, and represents a simple, cost-effective measure [7]. RDW is increasingly recognized as a marker for predicting the onset and progression of HF. In HF patients, anisocytosis may signify a homeostatic response to the disease, potentially indicating a link between ineffective erythropoiesis and chronic inflammation [8]. Traditionally, RDW has been underappreciated and primarily used to differentiate certain causes of anemia [9]. However, recent clinical evidence suggests that changes in RDW are associated with the development and adverse outcomes of stroke and cardiovascular disease [7,8,10,11]. Particularly in HF patients, RDW has been correlated with hospitalization rates and poor prognosis [12,13]. RDW is a readily applicable blood parameter for basic healthcare facilities, especially in Vietnam. Nonetheless, most existing evidence originates from regions such as the United States, Europe, Japan, and China, excluding Vietnam.

Therefore, this study aims to investigate RDW levels in hospitalized Vietnamese HF patients and assess the predictive ability of RDW for the severity of HF in this population.

## Materials and methods

### Study population

A cross-sectional study was conducted at Hue Central Hospital from February 2022 to February 2024, involving 351 patients categorized into two groups: those with HF and those without (see **Fig 1**). CHF and AHF were defined and classified based on the criteria outlined in the guidelines for the diagnosis and treatment of AHF and CHF of the European Society of Cardiology [14]. The control group consisted of patients with no history or symptoms of HF, LVEF ≥ 50%, and NT-proBNP < 125 pg/mL. Additionally, patients who had received blood transfusions or iron supplementation during hospitalization and those with a positive osmotic fragility test were excluded from the study. The project received approval from Hue Central Hospital and the Institutional Review Board of Duy Tan University (No: Đ23-24Y3-2) and adhered to the principles of the Declaration of Helsinki (2013 version).

**Fig 1.**
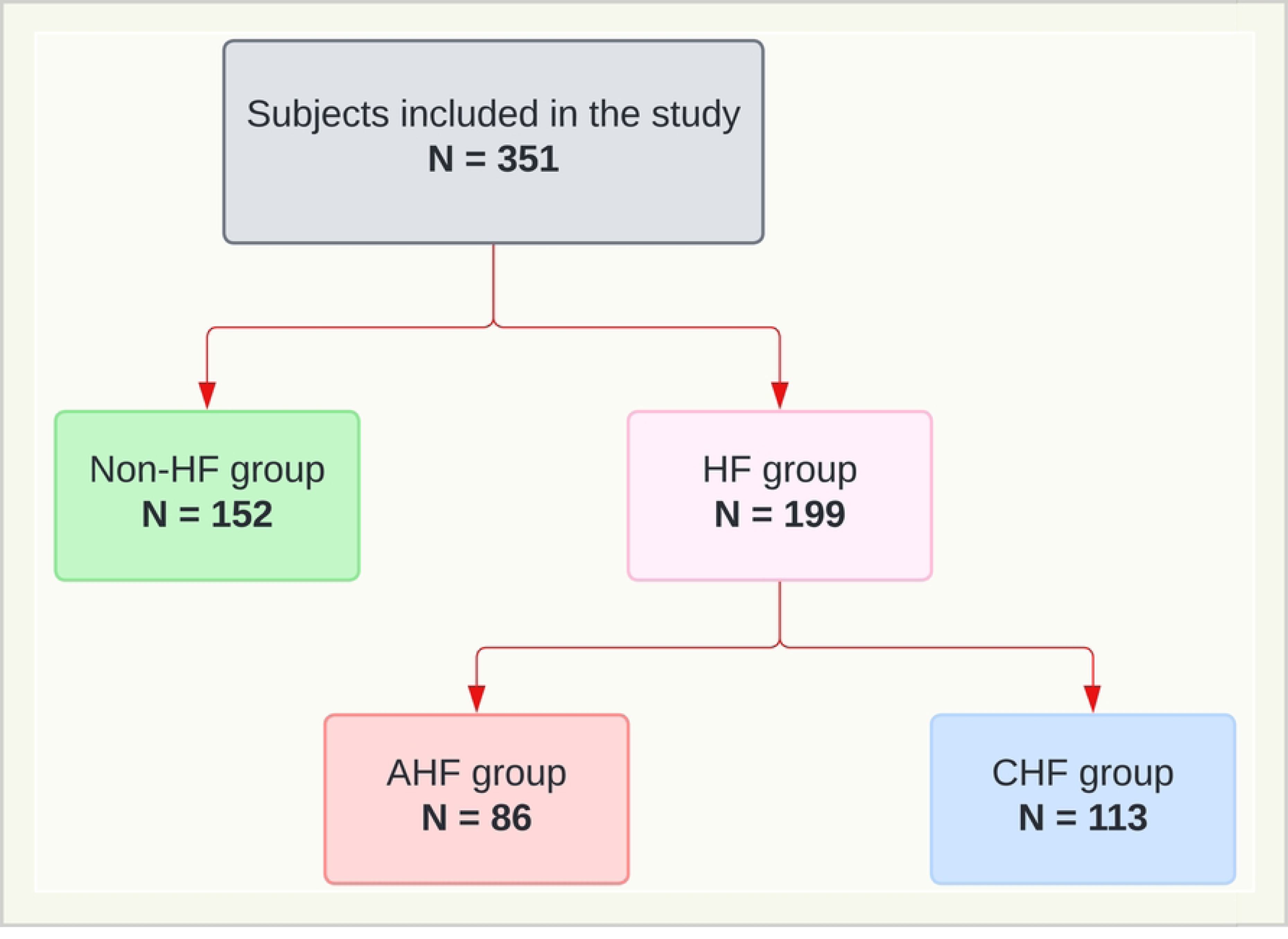
The research flowchart divides the research subjects into subgroups under the group. HF: heart failure; CHF: chronic heart failure; AHF: acute heart failure.

**Fig 2.**
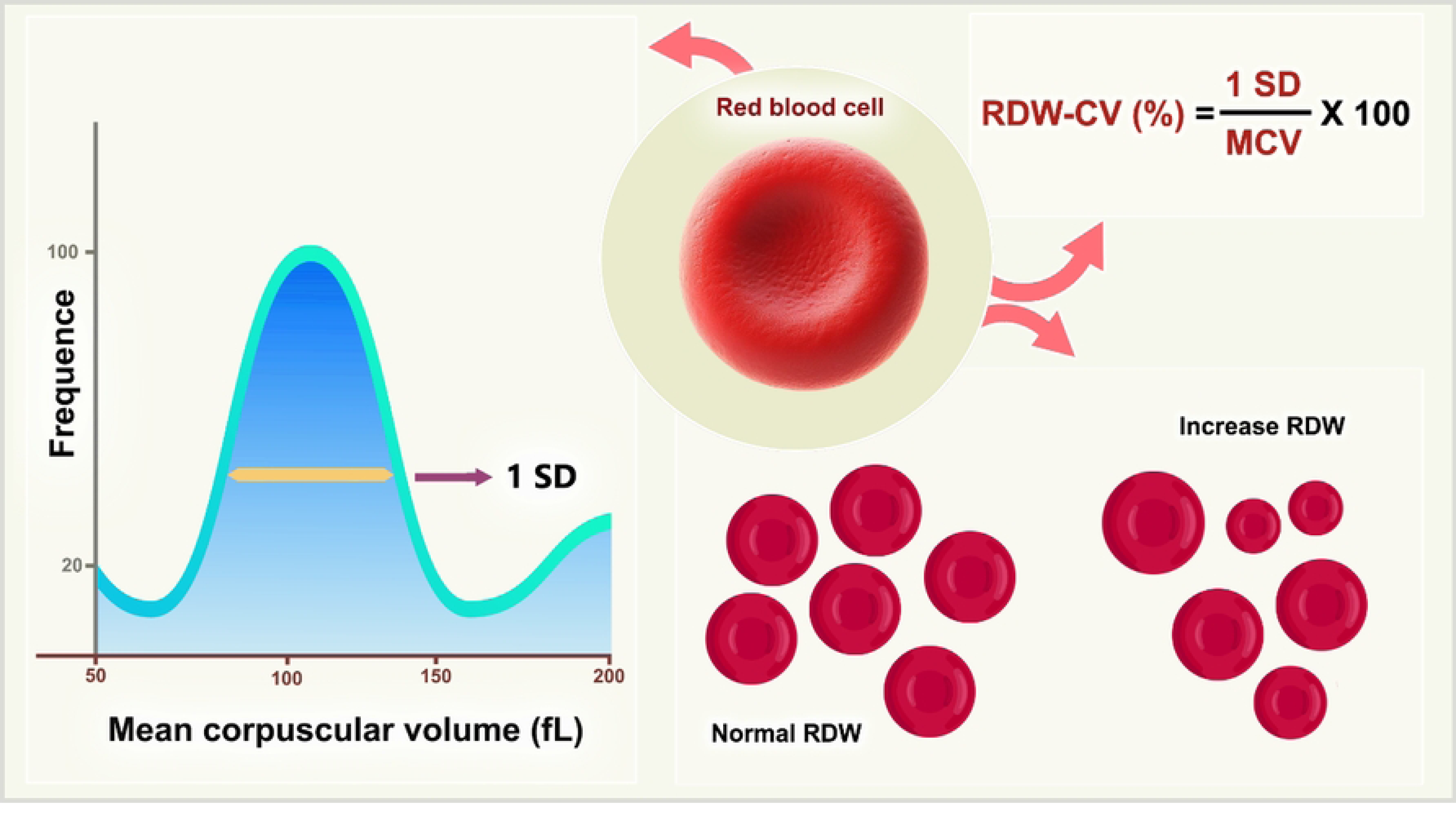
The RDW is found by the RDW-CV formula, 1 standard deviation divided by MCV. RDW-CV: red blood cell distribution width-coefficient of variation. RBC: red blood cell. SD: standard deviation. MCV: mean corpuscular volume.

### Data collection

The baseline evaluation of the patients encompassed demographic characteristics and clinical assessments, including medical history (hypertension, diabetes mellitus, dyslipidemia, coronary artery disease, chronic kidney disease, valvular heart disease, liver disease, and cardiac arrhythmias, as well as medication usage). Additionally, other clinical data such as body mass index (BMI), waist-hip ratio, heart rate, systolic blood pressure (SBP), diastolic blood pressure (DBP), and New York Heart Association classification (NYHA) were collected. These details were routinely collected upon admission and throughout hospitalization, utilizing a pre-established registry questionnaire.

The echocardiographic procedure adhered to the guidelines outlined by the American Society of Echocardiography for conducting a comprehensive transthoracic echocardiographic examination in adults, performed by expert echocardiographers [15].

The laboratory analyses were performed on fasting venous blood specimens and encompassed: brain natriuretic peptide, red blood cell count, hematocrit, hemoglobin, mean cell volume, mean cell hemoglobin content, mean cell hemoglobin concentration, platelet count, alanine aminotransferase, aspartate transaminase, glucose, urea, high-sensitivity troponin T, hemoglobin A1c, total cholesterol, triglyceride, low-density lipoprotein cholesterol, high-density lipoprotein cholesterol, non-high-density lipoprotein cholesterol, potassium level, sodium level, chloride level, and estimated glomerular filtration rate using CKD-EPI 2021 (eGFR, mL/min per 1.73 m^2^) [16].

### RDW measurement

Blood samples were collected from 2 mL of the peripheral vein using an ethylene diamine tetraacetic acid (EDTA) tube within 6 hours after admission and analyzed using the Sysmex XS-1000i automated hematology analyzer (Japan), located in the Hematology Department of Hue Central Hospital. RDW index was derived from the complete blood count obtained upon patients’ admission to the hospital. Two parameters of RDW were calculated to measure the extent of anisocytosis: the standard deviation (SD) and the coefficient variation (CV). RDW-CV is widely investigated and is calculated according to the following formula: RDW-CV = (SD of erythrocyte volume/mean corpuscular volume) × 100. The normal reference range for RDW in the hospital laboratories that participated in the study was 11.5 - 15% with an intra-assay variation of 2.38% and an inter-assay variation of 1% [13]. In this study, we reported RDW-CV and used RDW to represent it.

### Statistical analysis

All statistical analyses were performed using SPSS Version 26 (IBM, New York, United States), MedCalc Software Version 22.019 (MedCalc Software, Ostend, Belgium), and GraphPad Prism Version 10 (GraphPad Software, Boston, United States). Data normality was assessed using the Kolmogorov-Smirnov and Shapiro-Wilk tests. Normally distributed continuous variables were presented as mean ± standard deviation, while non-normally distributed variables were described as median values with interquartile ranges. Categorical variables were reported as frequencies and percentages. Fisher’s exact test was employed to assess intergroup differences in categorical variables, while differences in continuous variables were analyzed using either the unpaired T-test or the Mann-Whitney U test, as appropriate. Missing data were excluded from the analyses. Spearman’s correlation coefficient (r) and its corresponding p-value were calculated to explore the correlation between continuous variables. The area under the curve (AUC) was determined using the Wilson/Brown method to predict AHF. AUC comparison was conducted to evaluate the diagnostic performance of RDW compared to other biochemical tests, employing the Hanley and McNeil method [17]. Multivariable linear regression was used to evaluate the associations between clinical factors and left left atrium phasic strain and left atri left atrioventricular coupling index, respectively. Clinical confounding factors and variables that were significant (p < 0.05) in the univariate analysis were included in the multivariable model. To prevent multicollinearity among the univariate variables, a variance inflation factor of 5 was applied. Univariate logistic regression calculated odds ratios to predict HF. All statistical tests were two-sided, with a significance level of < 0.05.

## Results

### Baseline characteristics

The study involving 351 subjects revealed significant variations in blood cell count such as RBC, Hb, and RDW-CV between HF and non-HF patients. Detailed information regarding these indices is presented in **Table 1**.

**Table 1.** Baseline characteristics of the study groups.

| Characteristics | Non-HF (n= 152) | HF (n = 199) | p-value |
| --- | --- | --- | --- |
| **Baseline demographic and clinical features** |  |  |  |
| **Age (Years)** | 66.6 ± 12.1 | 70.1 ± 14.1 | 0.008 |
| **Female** | 115 (75.7) | 119 (59.8) | 0.002 |
| **Heart rate (bpm)** | 82 [76 - 90] | 81 [75 - 95] | 0.838 |
| **SBP (mmHg)** | 140 [130 - 170] | 130 [110 - 150] | $< 0.001$ |
| **DBP (mmHg)** | 80 [80 - 90] | 80 [70 - 80] | < 0.001 |
| **BMI (kg/m<sup>2</sup>)** | 22.4 [20.2 - 25.3] | 21.0 [19.2 - 23.1] | < 0.001 |
| **WHR** | 1.0 [0.9 – 1.0] | 1.0 [0.9 – 1.0] | 0.681 |
| **NYHA** |  | 2 [2 - 3] |  |
| **LVEF (%)** | 62.5 [61.0 - 64.3] | 55.0 [36.0 – 61.0] | < 0.001 |
| **Smoking** | 27.0 (17.8) | 31.0 (15.6) | 0.664 |
| **Alcohol** | 12.0 (7.9) | 16.0 (8.0) | 0.063 |
| **Diabetes** | 20.0 (13.2) | 51.0 (25.6) | 0.005 |
| **Dyslipidemia** | 39.0 (25.7) | 66.0 (33.2) | 0.158 |
| **Hypertension** | 116.0 (76.3) | 130 (65.3) | 0.034 |
| **Stent** | 3.0 (2.0) | 47.0 (23.6) | < 0.001 |
| **CABG** | 0.0 (0.0) | 11.0 (5.5) | 0.003 |
| **Atrial fibrillation** | 1.0 (0.7) | 39.0 (19.6) | < 0.001 |
| **Stroke** | 8.0 (5.3) | 15.0 (7.5) | 0.515 |
| **CKD** | 3.0 (2.0) | 21.0 (10.6) | 0.001 |
| **Medication usage** |  |  |  |
| **Antiplatelet** | 17.0 (11.2) | 25.0 (12.6) | 0.005 |
| **Beta blockers** | 24.0 (15.8) | 44.0 (22.1) | < 0.001 |
| **ACEi/ARB** | 81.0 (53.3) | 59.0 (29.7) | 0.299 |
| **MRA** | 4.0 (2.6) | 42.0 (21.1) | < 0.001 |
| **ARNI** | 0.0 (0.0) | 7.0 (3.5) | 0.001 |
| **SGLT2i** | 4.0 (2.6) | 26.0 (13.1) | < 0.001 |
| **Digoxin** | 0.0 (0.0) | 7.0 (3.5) | 0.001 |
| **Laboratory parameters** |  |  |  |
| **WBC (10<sup>9</sup>/L )** | 7.8 [6.5 - 9.6] | 7.6 [6.0 - 9.9] | 0.742 |
| **RBC (10<sup>12</sup>/L )** | 4.5 [4.2 - 4.7] | 3.8 [3.3 - 4.3] | < 0.001 |
| **Hb (g/dL)** | 13.1 [12.1 – 14.0] | 10.9 [8.7 - 12.5] | < 0.001 |
| **HCT (%)** | 39.5 [37.5 - 41.5] | 33.5 [27.3 - 38.2] | < 0.001 |
| **MCV (fL)** | 89.4 [86.1 - 92.6] | 88.3 [81.1 - 92.8] | 0.088 |
| **MCH (pg)** | 29.9 [28.8 - 30.9] | 29.0 [26.1 - 30.6] | 0.002 |
| **MCHC (g/dL)** | 33.2 [32.5 - 34.1] | 32.6 [31.7 - 33.4] | < 0.001 |
| **RDW-CV (%)** | 13.10 [12.23 - 13.78] | 14.90 [13.70 – 17.00] | < 0.001 |
| **PLT (10<sup>6</sup>/μL)** | 251 [215 - 294] | 230 [184 - 287] | 0.006 |
| **MPV (fL)** | 9.4 [8.3 - 9.9] | 8.7 [8.0 - 9.5] | 0.001 |
| **Glucose (mg/dL)** | 6.3 [5.3 - 7.8] | 6.4 [5.3 - 7.7] | 0.614 |
| **HbA1c (%)** | 6.3 [5.6 - 8.2] | 6.5 [5.8 - 8.1] | 0.634 |
| **AST (U/L)** | 24.0 [19.3 - 31.4] | 29.7 [21.5 - 41.2] | 0.002 |
| **ALT (U/L)** | 22.5 [15.4 - 33.2] | 22.0 [14.1 - 31.8] | 0.567 |
| **Cholesterol (mg/dL)** | 5.1 ± 1.1 | 4.4 ± 1.1 | < 0.001 |
| **Triglyceride (mg/dL)** | 1.5 [1.0 - 2.4] | 1.2 [0.9 - 1.8] | < 0.001 |
| **HDL-C (mg/dL)** | 1.3 ± 0.3 | 1.2 ± 0.3 | 0.013 |
| **Non-HDL-C (mg/dL)** | 3.8 ± 1.0 | 3.2 ± 1.0 | < 0.001 |
| **LDL-C (mg/dL)** | 3.2 [2.5 - 3.8] | 0.9 [0.6 - 2.4] | < 0.001 |
| **Urea (mg/dL)** | 4.7 [3.7 - 6.2] | 6.3 [5.0 - 8.6] | < 0.001 |
| **Creatinine (mg/dL)** | 67 [55 - 79] | 84 [61 - 121] | < 0.001 |
| **Potassium (mEq/L)** | 3.4 [3.2 - 3.8] | 3.7 [3.3 - 4.2] | < 0.001 |
| **Sodium (mEq/L)** | 138 [134 – 139] | 136 [134 - 140] | 0.278 |
| **Chloride (mEq/L)** | 101 [99 - 103] | 101 [97 - 105] | 0.478 |
| **hs-TnT (ng/L)** | 7.0 [6.0 – 11.0] | 18.5 [9.0 – 31.5] | < 0.001 |
| **NT-proBNP (pg/mL)** | 64 [32 - 99] | 1807 [487 - 8353] | < 0.001 |

Values are presented as mean ± standard deviation, median [25th interquartile - 75th interquartile], or number (%) as appropriate. SBP: systolic blood pressure; DBP: diastolic blood pressure; BMI: body mass index; WHR: waist hip ratio; NYHA: New York Heart Association; LVEF: left ventricular ejection fraction; CABG: coronary artery bypass graft; CKD: chronic kidney disease; ACEi/ARB: angiotensin-converting enzyme inhibitor/angiotensin receptor blocker; MRA: mineralocorticoid receptor antagonist; ARNI: angiotensin receptor neprilysin inhibitor; SGLT2i: sodium-glucose cotransporter 2 inhibitor; WBC: white blood cell; RBC: red blood cell; Hb: hemoglobin; HCT: hematocrit; MCV: mean corpuscular volume; MCH: mean corpuscular hemoglobin; MCHC: mean corpuscular hemoglobin concentration; RDW-CV: red blood cell distribution width coefficient of variation; PLT: platelet; MPV: mean platelet volume; HbA1c: hemoglobin A1c; AST: aspartate aminotransferase; ALT: alanine aminotransferase; HDL-C: high-density lipoprotein cholesterol; Non-HDL-C: non-high-density lipoprotein cholesterol; LDL-C: low-density lipoprotein cholesterol; hs-TnT: high-sensitivity troponin T; NT-proBNP: N-terminal pro-B-type natriuretic peptide.

### Comparison of RDW among different study groups

We observed significantly higher RDW-CV levels in AHF patients compared to the non-HF group. Additionally, HF patients with preserved ejection fraction had lower RDW-CV values compared to those with EF < 50%. Moreover, RDW levels increased with higher NYHA classifications. Detailed findings are illustrated in **Fig 3**.

**Fig 3.**
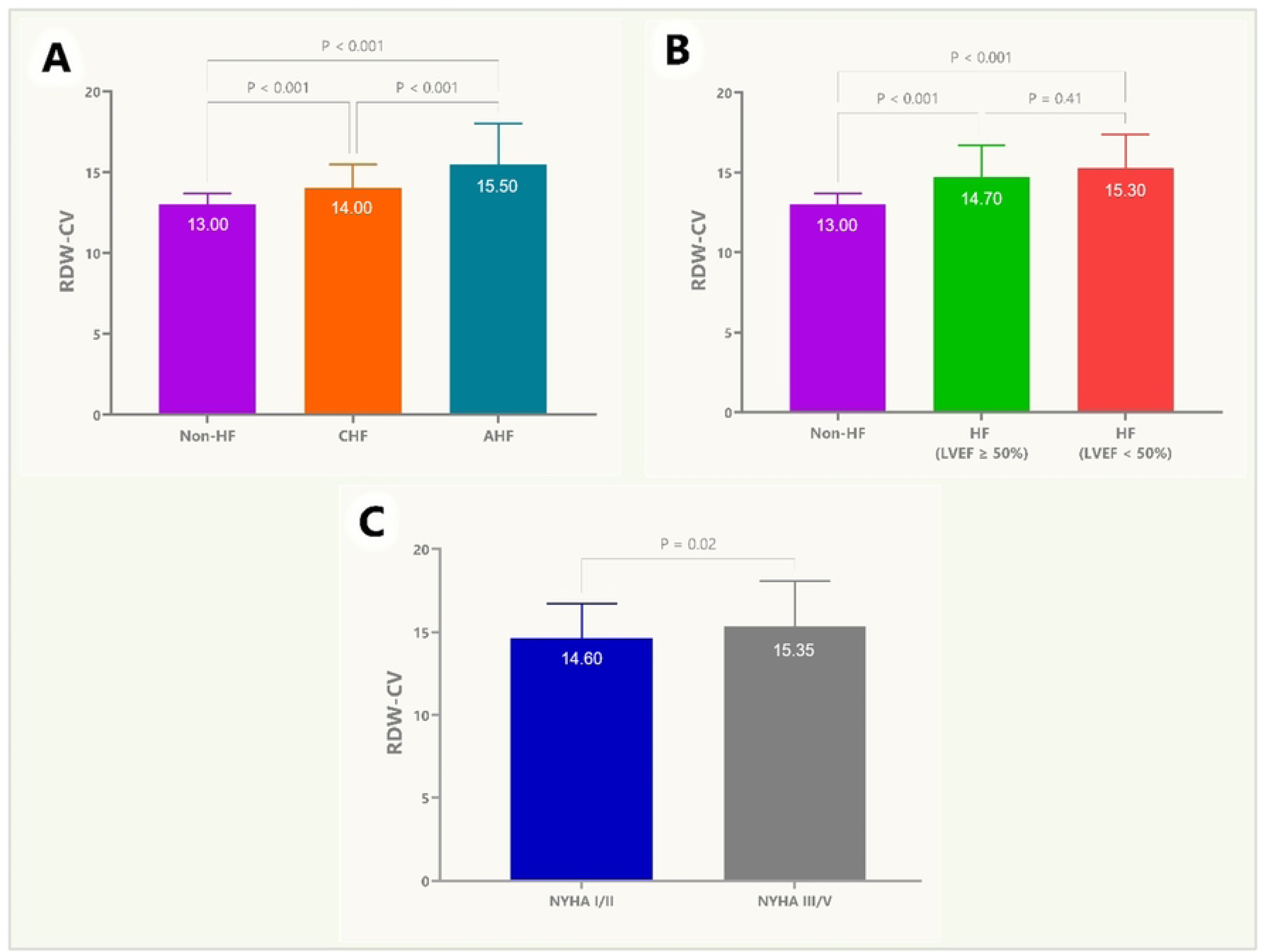
Comparison of RDW-CV among different study groups. (A) Illustrates the comparison of RDW-CV among Non-HF, CHF, and AHF groups. (B) Compares RDW-CV in HF patients with LVEF ≥ 50 and < 50. (C) Compares RDW-CV in NYHA I/II and NYHA III/IV groups. RDW-CV: red blood cell distribution width-coefficient of variation; HF: heart failure; CHF: chronic heart failure; AHF: acute heart failure; LVEF: left ventricular ejection fraction; NYHA: New York Heart Association.

### The predictive value of RDW for AHF

The prognostic significance of AHF in patients with CHF, as determined by RDW, is elucidated through ROC analysis, yielding an AUC of 0.651 (p < 0.001). The identified cutoff point stands at 13.85%, boasting a sensitivity of 86.05% and specificity of 36.94%. Comparative analysis between RDW and hs-TnT reveals a superior AUC for the latter. Nevertheless, employing the Hanley and McNeil method to discern the statistical significance between these AUCs, we find the discrepancy to be nonsignificant (p > 0.05). Conversely, RDW exhibits statistically significant differences (p < 0.05) when compared to WBC, PLT, and MCV. **Fig 4** further illustrates the predictive utility of AHF across various indices, including WBC, PLT, MCV, and hs-TnT.

**Fig 4.**
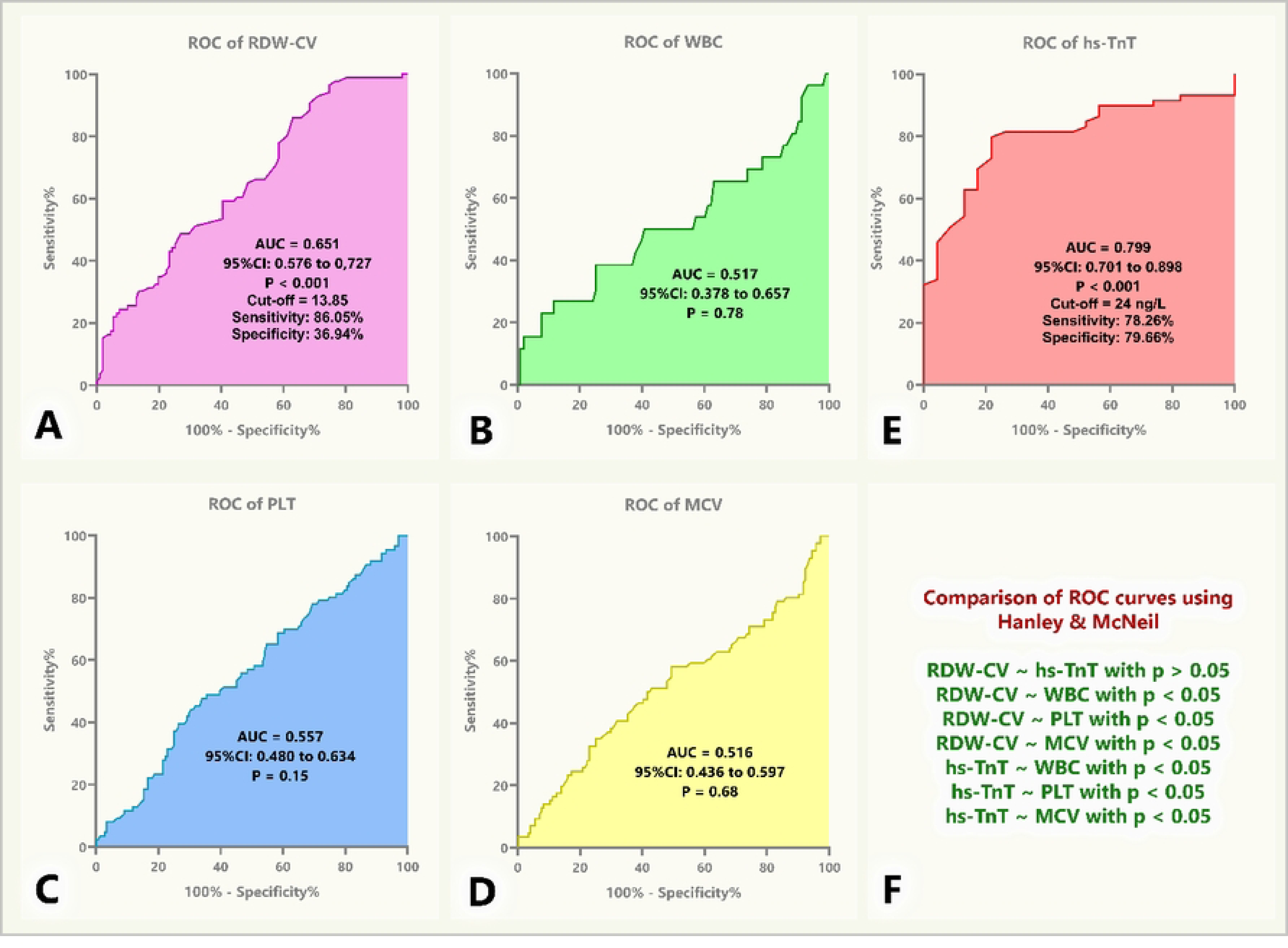
Receiver operating characteristic curves for predicting AHF among HF patients. (A-E) Depict the ROC curves of RDW-CV, WBC, PLT, MCV, and hs-TnT, respectively. (F) Compares the differences between the AUCs of RDW-CV, WBC, PLT, MCV, and hs-TnT using the Hanley & McNeil method. ROC: receiver operating characteristic; AUC: area under the curve; RDW-CV: red blood cell distribution width coefficient of variation; WBC: white blood cell; hs-TnT: high-sensitivity troponin T; PLT: platelet; MCV: mean corpuscular volume; AHF: acute heart failure.

### Correlation analysis of RDW and clinical, subclinical indices in HF patients

There is a positive correlation between the RDW-CV index and NT-proBNP and hs-TnT. Conversely, there is a negative correlation between the RDW-CV index and GFR. Furthermore, other parameters are clearly depicted in the heat map (see **Fig 5**).

**Fig 5.**
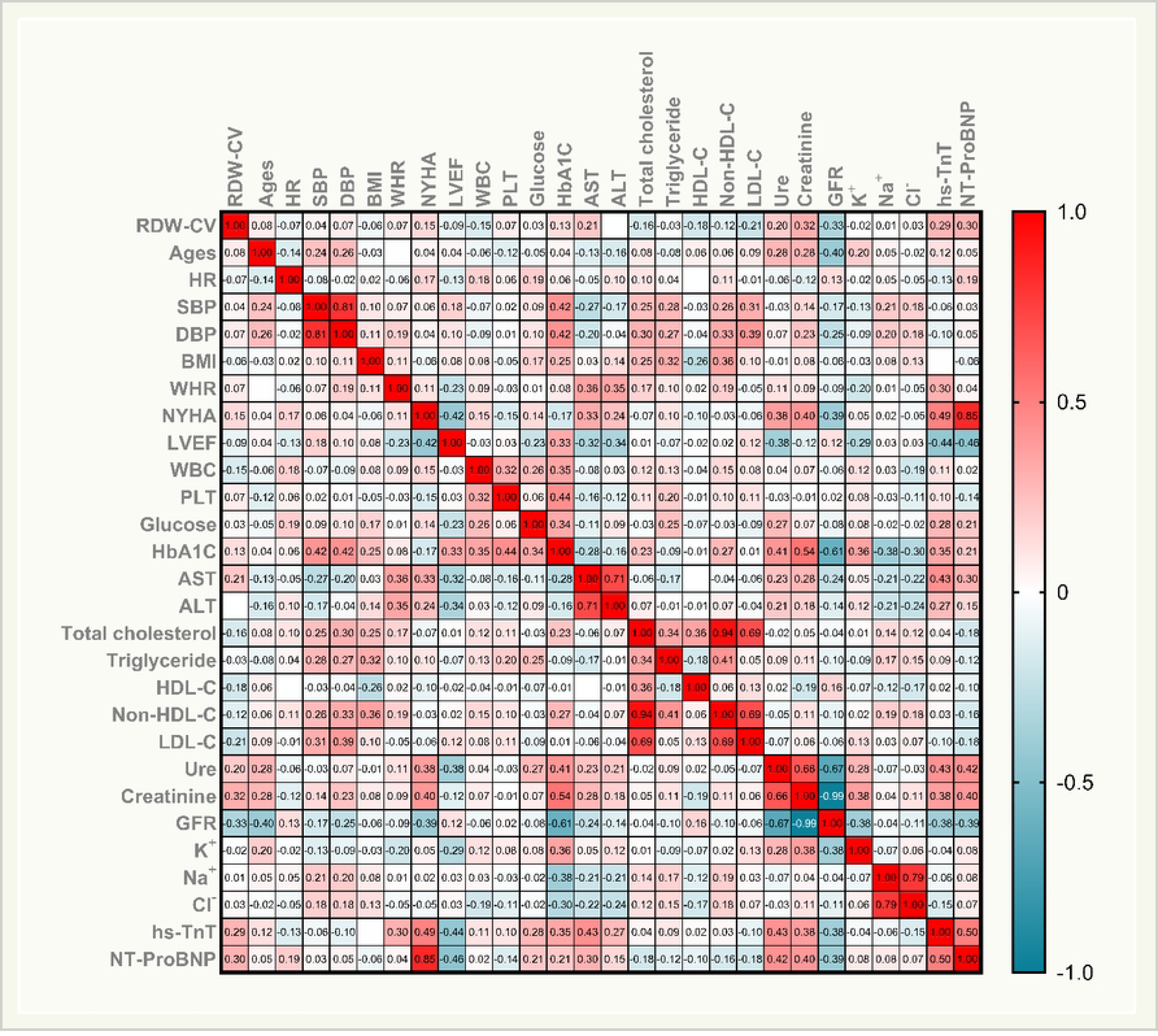
The heat map shows RDW-CV’s correlation with clinical and subclinical indices in HF patients. HR: heart rate; SBP: systolic blood pressure; DBP: diastolic blood pressure; BMI: body mass index; WHR: waist hip ratio; NYHA: New York Heart Association; LVEF: left ventricular ejection fraction; WBC: white blood cell; RDW-CV: red blood cell distribution width coefficient of variation; PLT: platelet; MPV: mean platelet volume; HbA1c: hemoglobin A1c; AST: aspartate aminotransferase; ALT: alanine aminotransferase; HDL-C: high-density lipoprotein cholesterol; Non-HDL-C: non-high-density lipoprotein cholesterol; LDL-C: low-density lipoprotein cholesterol; GFR: glomerular filtration rate, K^+^: Potassium; Na^+^: sodium; Cl^-^: chloride; hs-TnT: high-sensitivity troponin T; NT-proBNP: N-terminal pro-B-type natriuretic peptide.

### Linear and logistic regression analysis

**Table 2** illustrates the findings from both univariate and multivariate analyses conducted on RDW-CV concerning heart failure (HF). Univariate regression analysis unveiled significant associations between RDW-CV and several hematological parameters including RBC, Hb, HCT, MCV, MCH, MCHC, PLT, MPV, and GFR in HF patients. Multivariate analysis further elucidated associations between RDW-CV and Hb (β = 2.431, p = 0.040) as well as HCT (β = −3.355, p = 0.007) in this patient cohort. Additionally, **Fig 6** portrays the outcomes of logistic regression analyses for RDW-CV in HF patients, underscoring elevated RDW-CV levels as significant risk factors for HF.

**Fig 6.**
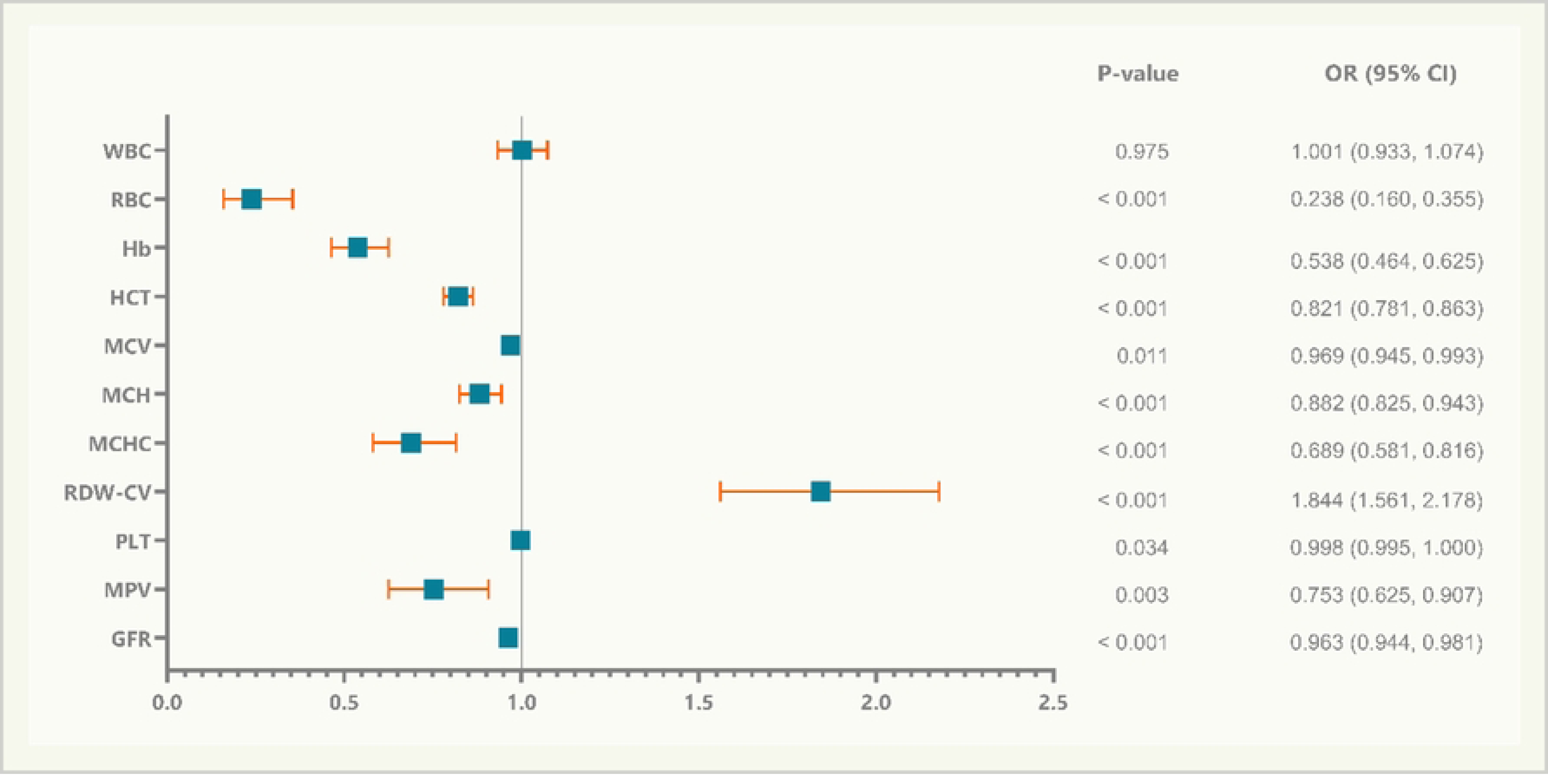
HF susceptibility was predicted via logistic regression using hematologic parameters and GFR as covariates. In the univariable analysis, elevated levels of RDW-CV along with a lower level of RBC and Hb, were identified as risk factors for HF. WBC: white blood cell; RBC: red blood cell; Hb: hemoglobin; HCT: hematocrit; MCV: mean corpuscular volume; MCH: mean corpuscular hemoglobin; MCHC: mean corpuscular hemoglobin concentration; RDW-CV: red blood cell distribution width coefficient of variation; PLT: platelet; MPV: mean platelet volume; GFR: glomerular filtration rate.

**Table 2.** Factors associated with increasing levels of RDW in linear regression.

| Factors | Univariable |  |  |  | Multivariable |  |  |  |
| --- | --- | --- | --- | --- | --- | --- | --- | --- |
| | $\beta$ | p-value | 95% CI | | $\beta$ | p-value | 95% CI | |
| **Sex** | 0.002 | 0.981 | -1.027 | 1.052 |  |  |  |  |
| **Age (years)** | -0.028 | 0.698 | -0.043 | 0.029 |  |  |  |  |
| **HR (beats/min)** | -0.082 | 0.253 | -0.04 | 0.011 |  |  |  |  |
| **BMI (kg/m<sup>2</sup>)** | -0.024 | 0.735 | -0.212 | 0.15 |  |  |  |  |
| **NYHA** | 0.118 | 0.099 | -0.098 | 1.128 |  |  |  |  |
| **Smoking** | -0.038 | 0.601 | -1.769 | 1.026 |  |  |  |  |
| **Alcohol** | 0.026 | 0.798 | -1.007 | 1.306 |  |  |  |  |
| **Diabetes** | -0.045 | 0.532 | -1.531 | 0.793 |  |  |  |  |
| **Dyslipidemia** | 0.100 | 0.161 | -0.308 | 1.848 |  |  |  |  |
| **Hypertension** | -0.009 | 0.903 | -1.138 | 1.005 |  |  |  |  |
| **Stent** | -0.048 | 0.501 | -1.602 | 0.786 |  |  |  |  |
| **CABG** | -0.022 | 0.759 | -2.570 | 1.878 |  |  |  |  |
| **Atrial fibrillation** | -0.031 | 0.666 | -1.190 | 0.762 |  |  |  |  |
| **Stroke** | -0.058 | 0.417 | -2.796 | 1.162 |  |  |  |  |
| **CKD** | -0.008 | 0.907 | -1.748 | 1.553 |  |  |  |  |
| **Antiplatelet** | -0.078 | 0.447 | -1.354 | 0.602 |  |  |  |  |
| **LVEF (%)** | -0.115 | 0.139 | -0.072 | 0.010 |  |  |  |  |
| **WBC (10<sup>9</sup>/L)** | -0.048 | 0.642 | -0.143 | 0.089 |  |  |  |  |
| **RBC (10<sup>12</sup>/L)** | -0.143 | **0.046** | -1.287 | -0.012 | 0.906 | 0.070 | -0.218 | 5.425 |
| **Hb (g/dL)** | -0.456 | **< 0.001** | -0.906 | -0.514 | 2.431 | **0.040** | 0.119 | 4.936 |
| **HCT (%)** | -0.405 | **< 0.001** | -0.283 | -0.146 | -3.355 | **0.007** | -2.111 | -0.343 |
| **MCV (fL)** | -0.446 | **< 0.001** | -0.202 | -0.113 | 0.950 | 0.512 | -0.450 | 0.895 |
| **MCH (pg)** | -0.476 | **< 0.001** | -0.552 | -0.324 | -0.507 | 0.777 | -2.513 | 1.884 |
| **MCHC (g/dL)** | -0.314 | **< 0.001** | -1.083 | -0.434 | -0.881 | 0.088 | -3.201 | 0.226 |
| **PLT (10<sup>9</sup>/L)** | 0.073 | 0.307 | -0.003 | 0.008 |  |  |  |  |
| **MPV (fL)** | -0.113 | 0.114 | -0.785 | 0.085 |  |  |  |  |
| **GFR**<br>**(mL/min/1.73m<sup>2</sup>)** | -0.262 | **0.010** | -0.059 | -0.008 | -0.143 | 0.053 | -0.037 | 0.000 |
| **NT-proBNP**<br>**(pg/mL)** | 0.097 | 0.176 | 0.000 | 0.000 |  |  |  |  |

Bold value indicates statistical significance at the p < 0.05 level. CI: confidence interval; HR: heart rate; SBP: systolic blood pressure; DBP: diastolic blood pressure; BMI: body mass index; WHR: waist hip ratio; NYHA: New York Heart Association; LVEF: left ventricular ejection fraction; CABG: coronary artery bypass graft; CKD: chronic kidney disease; ACEi/ARB: angiotensin-converting enzyme inhibitor/angiotensin receptor blocker; MRA: mineralocorticoid receptor antagonist; ARNI: angiotensin receptor neprilysin inhibitor; SGLT2i: sodium-glucose cotransporter 2 inhibitor; WBC: white blood cell; RBC: red blood cell; Hb: hemoglobin; HCT: hematocrit; MCV: mean corpuscular volume; MCH: mean corpuscular hemoglobin; MCHC: mean corpuscular hemoglobin concentration; RDW-CV: red blood cell distribution width coefficient of variation; PLT: platelet; MPV: mean platelet volume; GFR: glomerular filtration rate; NT-proBNP: N-terminal pro-B-type natriuretic peptide.

**Table 3** outlines independent risk factors identified in the complete blood count of AHF patients, where RBC and RDW-CV > 13.85% were recognized as independent risk factors with corresponding odds ratios of 0.313 (95% CI, 0.140-0.701; p = 0.005) and 2.644 (95% CI, 1.190-5.875; p = 0.017), respectively.

**Table 3.** Independent risk factors in complete blood count of AHF (analyzed by logistic regression model).

| Factors | Univariable |  |  |  | Multivariable |  |  |  |
| --- | --- | --- | --- | --- | --- | --- | --- | --- |
|  | OR<br>ratio | p-value | 95% CI |  | OR<br>ratio | p-value | 95% CI |  |
| **WBC (10<sup>9</sup>/L)** | 1.102 | 0.103 | 0.981 | 1.237 |  |  |  |  |
| **RBC (10<sup>12</sup>/L)** | 0.458 | **< 0.001** | 0.304 | 0.689 | 0.313 | **0.005** | 0.140 | 0.701 |
| **Hb (g/dL)** | 0.737 | **< 0.001** | 0.643 | 0.846 | 0.085 | 0.074 | 0.006 | 1.270 |
| **HCT (%)** | 0.915 | **< 0.001** | 0.874 | 0.957 | 2.294 | 0.068 | 0.940 | 5.595 |
| **MCV (fL)** | 0.998 | 0.877 | 0.971 | 1.026 |  |  |  |  |
| **MCH (pg)** | 0.966 | 0.339 | 0.899 | 1.037 |  |  |  |  |
| **MCHC (g/dL)** | 0.760 | **0.009** | 0.619 | 0.934 | 1.397 | 0.385 | 0.657 | 2.967 |
| **PLT (10<sup>9</sup>/L)** | 0.998 | 0.299 | 0.995 | 1.001 |  |  |  |  |
| **MPV (fL)** | 1.145 | 0.271 | 0.899 | 1.458 |  |  |  |  |
| **RDW-CV >**<br>**13.85%** | 3.612 | **< 0.001** | 1.755 | 7.432 | 2.644 | **0.017** | 1.190 | 5.875 |

Bold value indicates statistical significance at the p < 0.05 level. CI: confidence interval; OR: odd ratio; WBC: white blood cell; RBC: red blood cell; Hb: hemoglobin; HCT: hematocrit; MCV: mean corpuscular volume; MCH: mean corpuscular hemoglobin; MCHC: mean corpuscular hemoglobin concentration; RDW-CV: red blood cell distribution width coefficient of variation; PLT: platelet; MPV: mean platelet volume.

## Discussion

RDW is a simple, fast, cost-effective hematological parameter with results routinely provided in complete blood count. Our study aimed to compare RDW values between HF patients and the control group. Additionally, it evaluated the predictive value of RDW in anticipating the likelihood of HF exacerbation. The study was conducted on 351 subjects in Vietnam, divided into two groups: those with HF and those without.

### RDW in HF Patients

In our study, RDW levels in HF patients tended to be higher than in the non-HF group with p < 0.001 (**Fig 3**). Moreover, patients with elevated RDW levels were more likely to have HF compared to those with normal levels (**Fig 6**). Atac Celik et al. (2012) also found that the HF group had higher RDW compared to the control group [18]. Additionally, Yuxiang Dai et al. (2014) revealed that half of the HF patients had an RDW value above the upper limit of the normal range [19].

In our study, AHF patients had higher RDW levels compared to the CHF group and the control group (p < 0.001). Additionally, NYHA III/IV patients had higher RDW levels than NYHA I/II patients, with a significant difference observed (p = 0.02) (**Fig 3**). Similarly, Jaewon Oh et al. (2009) also found increased RDW levels in AHF patients [20].

Currently, the pathophysiological mechanism underlying the increased RDW in HF patients remains incompletely understood. Some recent literature suggests that inflammation, activation of the neuroendocrine system, and adrenergic activation in HF patients may influence the erythrocyte maturation process, leading to elevated RDW [13,21,22]. Red blood cells are formed in the bone marrow from erythroid colony-forming unit-erythroid progenitors and undergo maturation into mature erythrocytes through a series of developmental stages [23]. Disruption of various biological pathways, such as aging, inflammation, oxidative stress, nutritional deficiencies, impaired renal function, dyslipidemia, and alterations in RBC deformability or fragmentation, has been linked to impaired erythropoiesis, resulting in increased RDW [13,21,24–26]. In our study, variations in hematological parameters (RBC, Hb, HCT, MCV, MCH, and MCHC) were found to influence RDW outcomes. Particularly, Hb and HCT emerged as two parameters exerting independent effects on RDW results in the multivariable linear regression model (see **Table 2**).

Inflammation is recognized as a significant contributor to the pathophysiology of HF [27]. In HF, both cell-and cytokine-mediated inflammatory pathways are activated, causing bone marrow dysfunction and premature release of erythrocytes into the bloodstream. Inflammation inhibits bone marrow function and iron metabolism, while pro-inflammatory cytokines inhibit erythropoietin-induced erythrocyte maturation and proliferation, thus increasing RDW [28]. The correlation between RDW and inflammatory markers such as interleukin-6 (IL-6) and C-reactive protein (CRP) has been reported [13]. Pro-inflammatory cytokines like TNF-α, IL-1β, and IL-6 have been shown to reduce renal erythropoietin (EPO) synthesis, desensitize erythroid progenitor cells to EPO, and inhibit erythropoietin receptor expression, resulting in impaired erythroid progenitor cell proliferation, reduced RBC maturation, and increased RDW [29].

Neurohormonal activation is recognized as one of the primary mechanisms driving the advancement of HF, and therapeutic inhibition of neurohormonal systems has emerged as the fundamental approach in modern pharmacotherapy for HF [30]. Erythrocyte progenitor cells are stimulated by the activation of neurohumoral and adrenergic systems, leading to reduced erythropoiesis and consequently, elevated RDW levels. Consequently, irrespective of the underlying cause, the activation of adrenergic and neurohumoral systems directly contributes to the increase in RDW [18,31].

HF often coexists with several comorbidities, among which renal dysfunction holds particular importance. Cardiac and renal diseases intricately interact in both acute and chronic settings, forming a complex bidirectional relationship. Pathophysiologically, these conditions share common pathways, including inflammatory and direct cellular immune-mediated mechanisms, stress-mediated and neurohormonal responses, metabolic and nutritional alterations such as bone and mineral disorders, changes in hemodynamics and acid-base or fluid status, and the onset of anemia [32]. These mechanisms contribute to explaining the renal impairment in HF patients. Additionally, these mechanisms have also been implicated in increasing the RDW levels. In fact, our study observed an inverse correlation between RDW and Glomerular Filtration Rate (GFR) with r = −0.33 and p = 0.001, as depicted in **Fig 5**.

### Predictive Ability of RDW for AHF Onset in HF Patients

Previous studies have shown that an increase in RDW is associated with an increased risk of hospitalization due to AHF [33,34]. The study by Ferreira et al. (2013) demonstrated that high RDW at admission is a predictor of slower diuretic response [35]. This provides us with a positive insight into RDW levels in AHF patients.

In our study, RDW demonstrated predictive capability for AHF in HF patients with an AUC of 0.651, p < 0.001. The cutoff value of RDW in our study for predicting AHF was 13.85%, with a sensitivity of 86.05% and specificity of 36.94%. Furthermore, the predictive value of RDW for AHF was comparable to hs-TnT, with no significant difference in the AUC for predicting acute decompensated HF between these two parameters (**Fig 4**). Hs-TnT elevation is common among patients with AHF [36]. There is no doubt that an increased troponin level predicts outcomes in AHF, and the greater the elevation, the poorer the prognosis. It’s crucial to recognize that cardiac troponin indicates myocardial necrosis and is mainly employed for acute coronary syndrome diagnosis, which may precipitate an AHF episode. In this context, alterations in troponin levels over successive tests and the highest recorded level offer significant diagnostic and prognostic insights [37,38]. We found that a cost-effective and widely available test like RDW has an equivalent value to hs-TnT in predicting AHF.

Additionally, in our study, NT-proBNP exhibited a positive correlation with RDW in HF patients (**Fig 5**). There is no disputing the value of NT-proBNP in predicting and ruling out AHF. Numerous studies have demonstrated the excellent role of NT-proBNP in HF patients, especially in AHF cases [39–42]. However, in developing countries, the cost of performing NT-proBNP tests is prohibitively high and not available at primary healthcare facilities. Conversely, RDW is readily available and very inexpensive. Our study indicates that the positive correlation between RDW and NT-proBNP could open the door for the application of this index in primary healthcare facilities as well as in developing countries like Vietnam.

### Limitations of the study

Our study acknowledges several limitations. Firstly, relying solely on RDW without concurrent inflammatory markers such as C-reactive protein and gamma-glutamyl transferase may result in an incomplete assessment of inflammatory status, given RDW’s susceptibility to various conditions. Additionally, we lacked control over laboratory parameters linked to RDW variation and utilized a single blood analyzer machine (Sysmex XS-1000i, Japan) without inter-machine comparison. Despite efforts to mitigate selection bias through sequential sampling during routine HF visits, the initial conduct of our study at a single hospital with a cross-sectional approach may limit its generalizability. Moreover, the exclusivity of our study cohort to HF patients in central Vietnam further restricts generalization. Acknowledging the absence of serum iron ion data, a crucial confounder potentially affecting our results, future investigations will address this and consider the influence of unmeasured variables such as iron deficiency to enhance the study’s robustness.

## Conclusions

In the HF cohort, RDW levels were significantly higher compared to those in the non-HF cohort, and a positive correlation with NT-proBNP concentrations was observed. Additionally, RDW proved to be a valuable prognostic marker for AHF in HF patients. This study underscores the importance of further RDW research in HF to enhance understanding of its pathophysiology and improve risk assessment in HF management. Given its affordability and accessibility, RDW offers potential for application in resource-constrained environments.

## Data Availability

Data cannot be shared publicly due to certain local ethical constraints. Researchers who meet the criteria for access to confidential data may contact the author, Hai Nguyen Ngoc Dang (via email at).

## Acknowledgments

Nil

## References

1. Savarese G, Becher PM, Lund LH, Seferovic P, Rosano GMC, Coats AJS. Global burden of heart failure: a comprehensive and updated review of epidemiology. Cardiovascular Research. 2023;118: 3272–3287. doi:10.1093/cvr/cvac013

2. Lloyd-Jones DM, Larson MG, Leip EP, Beiser A, D’Agostino RB, Kannel WB, et al. Lifetime Risk for Developing Congestive Heart Failure: The Framingham Heart Study. Circulation. 2002;106: 3068–3072. doi:10.1161/01.CIR.0000039105.49749.6F

3. James SL, Abate D, Abate KH, Abay SM, Abbafati C, Abbasi N, et al. Global, regional, and national incidence, prevalence, and years lived with disability for 354 diseases and injuries for 195 countries and territories, 1990–2017: a systematic analysis for the Global Burden of Disease Study 2017. The Lancet. 2018;392: 1789–1858. doi:10.1016/S0140-6736(18)32279-7

4. Kittleson MM, Panjrath GS, Amancherla K, Davis LL, Deswal A, Dixon DL, et al. 2023 ACC Expert Consensus Decision Pathway on Management of Heart Failure With Preserved Ejection Fraction. Journal of the American College of Cardiology. 2023;81: 1835–1878. doi:10.1016/j.jacc.2023.03.393

5. Ji X, Ke W. Red blood cell distribution width and all-cause mortality in congestive heart failure patients: a retrospective cohort study based on the Mimic-III database. Front Cardiovasc Med. 2023;10: 1126718. doi:10.3389/fcvm.2023.1126718

6. Hullin R, Barras N, Abdurashidova T, Monney P, Regamey J. Red cell distribution width and prognosis in acute heart failure: ready for prime time! Intern Emerg Med. 2019;14: 195–197. doi:10.1007/s11739-018-1995-7

7. Diez-Silva M, Dao M, Han J, Lim C-T, Suresh S. Shape and Biomechanical Characteristics of Human Red Blood Cells in Health and Disease. MRS Bull. 2010;35: 382–388. doi:10.1557/mrs2010.571

8. Danese E, Lippi G, Montagnana M. Red blood cell distribution width and cardiovascular diseases. J Thorac Dis. 2015;7: E402–411. doi:10.3978/j.issn.2072Sdoi

9. Salvagno GL, Sanchis-Gomar F, Picanza A, Lippi G. Red blood cell distribution width: A simple parameter with multiple clinical applications. Critical Reviews in Clinical Laboratory Sciences. 2015;52: 86–105. doi:10.3109/10408363.2014.992064

10. Feng G-H, Li H-P, Li Q-L, Fu Y, Huang R-B. Red blood cell distribution width and ischaemic stroke. Stroke Vasc Neurol. 2017;2: 172–175. doi:10.1136/svn-2017-000071

11. Wang L, Wang C, Wu S, Li Y, Guo W, Liu M. Red blood cell distribution width is associated with mortality after acute ischemic stroke: a cohort study and systematic review. Ann Transl Med. 2020;8: 81–81. doi:10.21037/atm.2019.12.142

12. Gu F, Wu H, Jin X, Kong C, Zhao W. Association of red cell distribution width with the risk of 3-month readmission in patients with heart failure: A retrospective cohort study. Front Cardiovasc Med. 2023;10: 1123905. doi:10.3389/fcvm.2023.1123905

13. Xanthopoulos A, Giamouzis G, Dimos A, Skoularigki E, Starling R, Skoularigis J, et al. Red Blood Cell Distribution Width in Heart Failure: Pathophysiology, Prognostic Role, Controversies and Dilemmas. JCM. 2022;11: 1951. doi:10.3390/jcm11071951

14. McDonagh TA, Metra M, Adamo M, Gardner RS, Baumbach A, Böhm M, et al. 2023 Focused Update of the 2021 ESC Guidelines for the diagnosis and treatment of acute and chronic heart failure. European Heart Journal. 2023;44: 3627–3639. doi:10.1093/eurheartj/ehad195

15. Mitchell C, Rahko PS, Blauwet LA, Canaday B, Finstuen JA, Foster MC, et al. Guidelines for Performing a Comprehensive Transthoracic Echocardiographic Examination in Adults: Recommendations from the American Society of Echocardiography. Journal of the American Society of Echocardiography. 2019;32: 1–64. doi:10.1016/j.echo.2018.06.004

16. Maynard RD, Korpi-Steiner N, Cotten SW. Concordance of Chronic Kidney Disease Stage and Metformin Management Using CKD-EPI 2021 Race-Free Equation vs CKD-EPI 2009 Equation to Estimate Glomerular Filtration Rate. Clinical Chemistry. 2023;69: 202–204. doi:10.1093/clinchem/hvac195

17. Hanley JA, McNeil BJ. A method of comparing the areas under receiver operating characteristic curves derived from the same cases. Radiology. 1983;148: 839–843. doi:10.1148/radiology.148.3.6878708

18. Celik A, Koc F, Kadi H, Ceyhan K, Erkorkmaz U, Burucu T, et al. Relationship between red cell distribution width and echocardiographic parameters in patients with diastolic heart failure. The Kaohsiung J of Med Scie. 2012;28: 165–172. doi:10.1016/j.kjms.2011.06.024

19. Dai Y, Konishi H, Takagi A, Miyauchi K, Daida H. Red cell distribution width predicts short- and long-term outcomes of acute congestive heart failure more effectively than hemoglobin. Experimental and Therapeutic Medicine. 2014;8: 600– 606. doi:10.3892/etm.2014.1755

20. Oh J, Kang S-M, Hong N, Choi J-W, Lee S-H, Park S, et al. Relation Between Red Cell Distribution Width With Echocardiographic Parameters in Patients With Acute Heart Failure. Journal of Cardiac Failure. 2009;15: 517–522. doi:10.1016/j.cardfail.2009.01.002

21. Lippi G, Turcato G, Cervellin G, Sanchis-Gomar F. Red blood cell distribution width in heart failure: A narrative review. WJC. 2018;10: 6. doi:10.4330/wjc.v10.i2.6

22. Bekler A, Tenekecioglu E, Erbag G, Temiz A, Altun B, Barutcu A, et al. Relationship between red cell distribution width and long-term mortality in patients with non-ST elevation acute coronary syndrome. Anatol J Cardiol. 2015;15: 634–639. doi:10.5152/akd.2014.5645

23. Hattangadi SM, Wong P, Zhang L, Flygare J, Lodish HF. From stem cell to red cell: regulation of erythropoiesis at multiple levels by multiple proteins, RNAs, and chromatin modifications. Blood. 2011;118: 6258–6268. doi:10.1182/blood-2011-07-356006

24. Ananthaseshan S, Bojakowski K, Sacharczuk M, Poznanski P, Skiba DS, Prahl Wittberg L, et al. Red blood cell distribution width is associated with increased interactions of blood cells with vascular wall. Sci Rep. 2022;12: 13676. doi:10.1038/s41598-022-17847-z

25. Miglio A, Valente C, Guglielmini C. Red Blood Cell Distribution Width as a Novel Parameter in Canine Disorders: Literature Review and Future Prospective. Animals. 2023;13: 985. doi:10.3390/ani13060985

26. Joosse H-J, Van Oirschot BA, Kooijmans SAA, Hoefer IE, Van Wijk RAH, Huisman A, et al. In-vitro and in-silico evidence for oxidative stress as drivers for RDW. Sci Rep. 2023;13: 9223. doi:10.1038/s41598-023-36514-5

27. Reina-Couto M, Pereira-Terra P, Quelhas-Santos J, Silva-Pereira C, Albino-Teixeira A, Sousa T. Inflammation in Human Heart Failure: Major Mediators and Therapeutic Targets. Front Physiol. 2021;12: 746494. doi:10.3389/fphys.2021.746494

28. Khedar M, Sharma DK, Ola V. Red blood cell distribution width - A novel marker of inflammation and predictor of complications and outcomes among surgically managed patients. Formosan Journal of Surgery. 2021;54: 130–134. doi:10.4103/fjs.fjs_109_20

29. Förhécz Z, Gombos T, Borgulya G, Pozsonyi Z, Prohászka Z, Jánoskuti L. Red cell distribution width in heart failure: Prediction of clinical events and relationship with markers of ineffective erythropoiesis, inflammation, renal function, and nutritional state. American Heart Journal. 2009;158: 659–666. doi:10.1016/j.ahj.2009.07.024

30. Hartupee J, Mann DL. Neurohormonal activation in heart failure with reduced ejection fraction. Nat Rev Cardiol. 2017;14: 30–38. doi:10.1038/nrcardio.2016.163

31. Contreras Gutiérrez VH. Red cell distribution width: A marker of in-hospital mortality in ST-segment elevation myocardial infarction patients? Revista Médica del Hospital General de México. 2017;80: 165–169. doi:10.1016/j.hgmx.2016.10.001

32. Schefold JC, Filippatos G, Hasenfuss G, Anker SD, Von Haehling S. Heart failure and kidney dysfunction: epidemiology, mechanisms and management. Nat Rev Nephrol. 2016;12: 610–623. doi:10.1038/nrneph.2016.113

33. Felker GM, Allen LA, Pocock SJ, Shaw LK, McMurray JJV, Pfeffer MA, et al. Red Cell Distribution Width as a Novel Prognostic Marker in Heart Failure. Journal of the American College of Cardiology. 2007;50: 40–47. doi:10.1016/j.jacc.2007.02.067

34. Allen LA, Felker GM, Mehra MR, Chiong JR, Dunlap SH, Ghali JK, et al. Validation and Potential Mechanisms of Red Cell Distribution Width as a Prognostic Marker in Heart Failure. Journal of Cardiac Failure. 2010;16: 230–238. doi:10.1016/j.cardfail.2009.11.003

35. Ferreira JP, Santos M, Almeida S, Marques I, Bettencourt P, Carvalho H. Tailoring diuretic therapy in acute heart failure: insight into early diuretic response predictors. Clin Res Cardiol. 2013;102: 745–753. doi:10.1007/s00392-013-0588-8

36. Newby LK. High-Sensitivity Troponin in Acute Heart Failure Triage: Tacit but Not Confirmed. Circ: Heart Failure. 2019;12: e006241. doi:10.1161/CIRCHEARTFAILURE.119.006241

37. Wettersten N, University of California San Diego, Maisel A, Veterans Affairs San Diego Healthcare System, La Jolla, CA, USA. Role of Cardiac Troponin Levels in Acute Heart Failure. Cardiac Failure Review. 2015;1: 102. doi:10.15420/cfr.2015.1.2.102

38. Ledwoch J, Kraxenberger J, Krauth A, Schneider A, Leidgschwendner K, Schneider V, et al. Prognostic impact of high-sensitive troponin on 30-day mortality in patients with acute heart failure and different classes of left ventricular ejection fraction. Heart Vessels. 2022;37: 1195–1202. doi:10.1007/s00380-022-02026-x

39. Richards AM, Christchurch Heart Institute, University of Otago, Christchurch, New Zealand, Cardiovascular Research Institute, National University of Singapore, Singapore. Biomarkers In Acute Heart Failure — Cardiac And Kidney. Cardiac Failure Review. 2015;1: 107. doi:10.15420/cfr.2015.1.2.107

40. Adamo M, Pagnesi M, Mebazaa A, Davison B, Edwards C, Tomasoni D, et al. NT-proBNP and high intensity care for acute heart failure: the STRONG-HF trial. European Heart Journal. 2023;44: 2947–2962. doi:10.1093/eurheartj/ehad335

41. Singhal AK, Singh G, Singh S, Karunanand B, Agrawal S. Role of Pro-BNP in predicting outcome in acute heart failure patient presenting to a medical emergency: An observational study from North India. Journal of Family Medicine and Primary Care. 2023;12: 3156–3159. doi:10.4103/jfmpc.jfmpc_853_23

42. Boehmer JP, Nair DG, Wen G, An Q, Thakur PH, Gardner RS. HeartLogic Performs as Well as NT-proBNP to Rule out Acute Heart Failure at Point of Care. Journal of Cardiac Failure. 2019;25: S17–S18. doi:10.1016/j.cardfail.2019.07.052

